# Lightweight Model For The Prediction of COVID-19 Through The Detection And Segmentation of Lesions in Chest CT Scans

**DOI:** 10.1101/2020.10.30.20223586

**Authors:** Aram Ter-Sarkisov

## Abstract

We introduce a lightweight Mask R-CNN model that segments areas with the Ground Glass Opacity and Consolidation in chest CT scans. The model uses truncated ResNet18 and ResNet34 nets with a single layer of Feature Pyramid Network as a backbone net, thus substantially reducing the number of the parameters and the training time compared to similar solutions using deeper networks. Without any data balancing and manipulations, and using only a small fraction of the training data, COVID-CT-Mask-Net classification model with 6.12M total and 600K trainable parameters derived from Mask R-CNN, achieves **91**.**35**% COVID-19 sensitivity, **91**.**63**% Common Pneumonia sensitivity, **96**.**98**% true negative rate and **93**.**95**% overall accuracy on COVIDx-CT dataset (21191 images). We also present a thorough analysis of the regional features critical to the correct classification of the image. The full source code, models and pretrained weights are available on https://github.com/AlexTS1980/COVID-CT-Mask-Net.

## 1 Introduction

Most Deep Learning algorithms predicting COVID-19 from chest CT scans use one of the three approaches to classification: general-purpose feature extractor such as ResNet or DenseNet, or a specialized one, like COVIDNet-CT mapping the input to the predicted class, [GWW20, BGCB20, LQX^+^20, YWR^+^20, SZL^+^], a combination of feature extraction and a semantic segmentation/image mask, [JWX^+^20, WGM^+^20, ZZHX20] and a combination of regional instance extraction and global (image) classification, [TS20a, TS20b].

Each approach has certain drawbacks regardless of the achieved accuracy of the model. These drawbacks include a small size of the dataset [BGCB20], limited scope (only two classes: COVID-19 and Common Pneumonia (CP), COVID-19 and Control, COVID-19 and non-COVID-19), [SZL^+^, ZZHX20], large training data requirement [GWW20], large model size [LQX^+^20, TS20a]. In [TS20a] the drawback of using a large amount of data was addressed by training a Mask R-CNN [HZRS16] model to segment areas with lesions in chest CT scans. Then, the model was augmented with a classification head that predicts the class of the image. This allowed for using a much smaller dataset for training than, e.g. [GWW20] at the cost of the size of the model, which has 34.14M total and 2.45M trainable parameters.

In this paper we overcome this drawback by attempting several variants of two different backbone models, ResNet18 and ReNet34 [HZRS16] with a single Feature Pyramid Network (FPN) layer connected to the last backbone layer. The sizes of models vary from 4.02M to 24.63M parameters (segmentation model) and 4.25M to 24.86M (classification model), with only 0.6M trainable parameters in the classification model in [TS20a]. Segmentation models with a truncated ResNet34+FPN backbone (last block of layers deleted) with 11.74M parameters achieved a mean average precision (mAP) of 0.4476, which is at par with the top 25 results of MS COCO segmentation leaderboard, https://cocodataset.org/#detection-leaderboard. The classification model using this backbone, with 11.74M total parameters, of which only 0.6M are trainable, achieved a 91.76% COVID-19 sensitivity and 92.89% overall accuracy. An even smaller model, truncated ResNet18+FPN (6.12M parameters in the segmentation model and 6.35M in the classification model, of which also 0.6M are trainable) achieved mAP of 0.3932, COVID-19 sensitivty of 91.35% and overall accuracy of 93.95%.

## 2 Data and Models

We use the same datasets and train/validation/test splits as in [TS20a, TS20b] for a fair comparison. The raw chest CT scan data is taken from CNCB-COVID repository, [ZLS^+^20], http://ncov-ai.big.ac.cn/download. For the segmentation problem, the train/validation split is 500/150. All results reported in Table 2 were obtained on the validation split. The train/validation/test splits for the classification model are taken from COVIDx-CT [GWW20]: 3000 images were sampled randomly from the train split (over 60000 images) and used to train all COVID-CT-Mask-Net classifiers. Validation and test splits were used in full (21036 and 21192 images resp.).

Apart from the subtraction of the global mean and division by the global standard deviation, no other data manipulations were applied to either dataset.

The main contribution of this paper is the training of the lightweight segmentation and classification models with ResNet18+FPN and ResNet34+FPN backbones to produce results that beat or approach those of the full-sized ResNet50+FPN models with 4 FPN layers for both tasks. In all backbone nets the last (problem-specific) fully connected and average pooling layers were removed. For the full list of model sizes and comparison to the benchmarks, see Table 1. We consider three versions of each model:

**Table 1:**
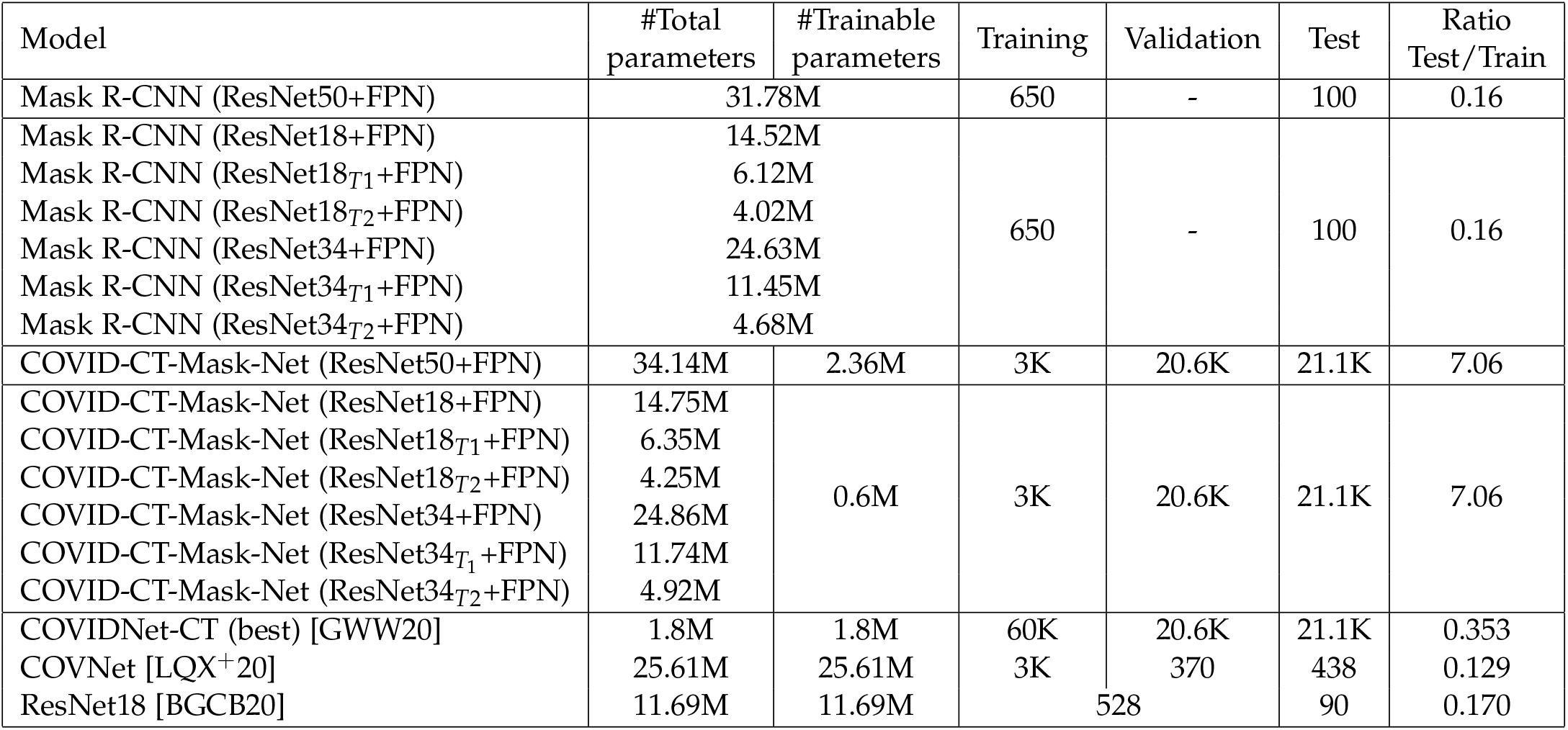
Comparison of the models’ sizes and data splits used for training, validation and testing. *T*1 and *T*2 refer to the truncated models (1 and 2), see Figure 1a. FPN is used in all our models because it helps with the reduction in the total number of parameters and improves the final result. The number of trainable parameters in the classifiers with ResNet18 and ResNet34 backbones varies insignificantly.

1. Full model. This is the baseline for each experiment, in Figure 1a it is the model that contains all blocks (green), and FPN module is connected to the last fourth block. FPN input is downsized from 512 to 256 maps.
2. ResNet 18/34_*T*1_: the first truncated model. The last ResNet block is removed, FPN is connected to Block 3, and FPN has the same number of maps (256) as the last block in ResNet.
3. ResNet 18/34_*T*2_: the second truncated model. The last two blocks in ResNet are removed, and FPN is connected to ResNet Block 2. FPN upsizes the input from 128 to 256 maps.

For the training and evaluation of the segmentation model we used only one positive class, ‘Lesion’, obtained by merging the masks for the Ground Glass Opacity (GGO) and Consolidation (C) areas, see [TS20b]. For the training and evaluation of the classification model, we use the labeling convention from COVIDx-CT and CNCB: 0 for the Control class, 1 for Common Pneumonia and 2 for COVID-19.

**Figure 1:**
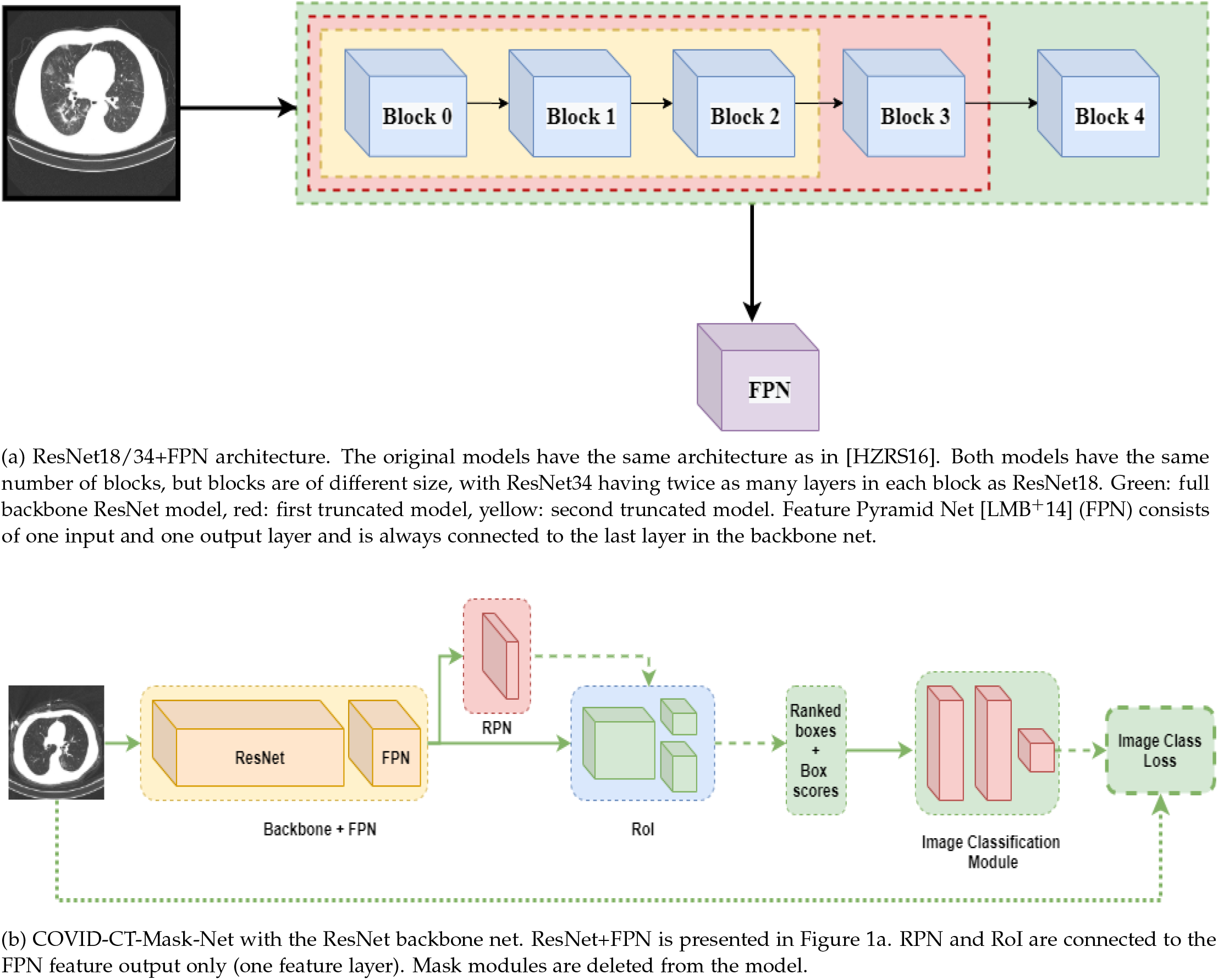
Architecture of the backbone nets (Figure 1a) and the lightweight COVID-CT-Mask-Net (Figure 1b). Best viewed in color.

## 3 Experimental results

For the explanation of the accuracy metrics and comparison, see [LMB^+^14], as we adapt MS COCO’s average precision (AP) at two Intersect over Union (IoU) threshold values and mean AP across 10 IoU thresholds between 0.5 and 0.95 with at 0.05 step. To test the models we used RoI and RPN NMS threshold of 0.75 and confidence score threshold of 0.75. The hyperparameters of the classification model are the same as in the best model in [TS20b], with the NMS threshold of 0.75 and RoI score_*θ*_ = −0.01, except that we reduce the RoI batch size from 256 to 128 and the total model size from 34.14M and the number of trainable parameters from 2.45M (ResNet50+FPN) to 6.12M and 0.6M respectively (ResNet18_*T*1_+FPN) with only about 2% drop in the COVID-19 sensitivity and 1.5% drop in overall accuracy. For the comparison to larger models, see [TS20b].

Results for training full and truncated lightweight models are presented in Table 2. The best segmentation model we trained, ResNet34 with a deleted last block (ResNet34_*T*1_+FPN) with 11.45M parameters achieves mAP of 44.76% and marginally outperforms the best model in [TS20b], ResNet50+FPN with merged masks, which is almost 3 times larger. The classification model derived from it also achieves the highest COVID-19 sensitivity among the lightweight models, 91.76%. The second-best segmentation model, ResNet18_*T*1_+FPN, achieves 0.4269 overall accuracy, with only 6.12M parameters. The classification model derived from it achieves the highest overall accuracy of 93.95% and the second-best COVID-19 sensitivity among the lightweight models of 91.35%. High segmentation performance does not immediately translate into the equally strong advantage in classification, but overall the models that did best for the segmentation task also achieved the highest accuracy in COVID-19 sensitivity, overall accuracy and true negative rate.

We experimented with a number of additional hacks for each model:

1. Replacement of SOFTMAX with SIGMOID activation function for the outputs of RoIs (segmentation model, test stage). Faster R-CNN implementation [RHGS15] uses SOFTMAX for scoring *C* outputs of each RoI (*C*:total number of classes, including background). The score of each non-background prediction is compared to the score threshold (RoI score_*θ*_ = 0.75) to decide whether to keep this prediction or discard, so obviously it is very unlikely to get more than a single prediction out of each RoI. At the same time, even low-ranking predictions are tested for Non-max suppression (0.75 in all models). Replacing SOFTMAX with SIGMOID makes predictions independent of each other in each RoI, and hence have a higher chance of being accepted as a prediction. This approach did not yield a consistent improvement across all models, so we left it out of the final result.
2. Removal of empty boxes/replication of the predictions (classification model). Deletion of empty boxes (bounding boxes with the area of 0) improved the models’ predictive power, but reduced the output size of the pre-defined RoI batch size (128), which is converted to a feature vector in the classification module **S**, and hence must remain fixed (see [TS20a] for details of batch to feature method). To resolve the problem, we applied a hack at this stage: the missing predictions (difference between the pre-defined RoI batch size and the current output) are sampled from the valid predictions *maintaining their ranking order*. What this means is that each sampled prediction is inserted in the batch between the box selected for replication and the next prediction. For example, if the predictions are [3, 1, 2] and the first and the last ones are sampled for replication, the batch becomes [3, 3, 1, 2, 2]. This maintains the order of ranking of the predictions in the sample, which is what the classifier learns to predict the class of the input image.
3. Removal of small areas in the data (segmentation model). Most areas with GGO and C are small, see [ZZX^+^20, ZYW^+^20, TS20a] for the detailed discussion of the distribution of lesions in chest CT scans. Training the segmentation model to predict small lesion areas leads both to lower precision at test stage, and lower COVID-19 sensitivity of the classification model. We decided to merge all GGO and C patches of less than 100 pixels with the background. As a result, the model’s accuracy improved, as the predictions were not biased towards very small areas.

### 3.1 Identification of areas critical for COVID-19 prediction

Apart from the CT scan segmentation and classification, deep learning models can help explain factors associated with COVID-19, e.g. in the form of attention maps [YMK^+^20, YWR^+^20] or using specialized tools like GSInquire [GWW20] that identify critical factors in CT scans. The advantage of using instance segmentation models like Mask R-CNN is the detection, scoring and segmentation of isolated areas (instances) that contribute to the condition (class of the image). This is a more accurate and explicit approach than either feature maps in vanilla convnets, that merely indicate the strength of presence of nameless features, or full-image pixel-level score maps in FCNs, that do not distinguish between different instances of the objects belonging to the same class. Mask R-CNN independently evolves separate instances of regional predictions that can overlap, both at bounding box and mask level.

This is illustrated in Figure 2 for the output of ResNet34_*T*1_ model. Figures 2a and 2b are COVID-19 positive, Figures 2c and 2d are COVID-19 negative (Control, no lesions at slice level), which is reflected in column 3 (column 3: no lesion mask). The first column is the input image overlaid with bounding box predictions for the lesion areas with a box confidence score and mask predictions for the object in the bounding box. Mask predictions are usually normalized using SIGMOID function, with a threshold of 0.5 that serves as a filter for the foreground (i.e. all pixels with scores exceeding the threshold are considered foreground/instance), but for the (combined) mask score map in column 2 in Figure 2, we used raw (before SIGMOID normalization) scores from Mask R-CNN. Each prediction is done by Mask R-CNN *independently*, i.e. the full path of extracting the RoI from the FPN layer using RoIAlign ([HGDG17]), predicting bounding box coordinates, filtering it through the de-convolution layer to obtain a fixed-size (28*×* 28) mask score map with pixel logits that is then resized to the size of the bounding box prediction is done independently for each object. Looking at the combined mask score maps, it becomes clear how COVID-CT-Mask-Net learns to use the score information. Each score map for the negative images contains only one prediction with a very low confidence score (*<* 0.01), for which COVID-CT-Mask-Net outputs large logit values for Class 0 (Figure 2, column 4). Score maps for COVID-19 images contain a number of large high-scoring predictions. The total number of predictions in each image is the same due to the RoI score_*θ*_ = − 0.01, we plotted only a small number of the highest-scoring RoIs to avoid image cluttering.

**Figure 2:**
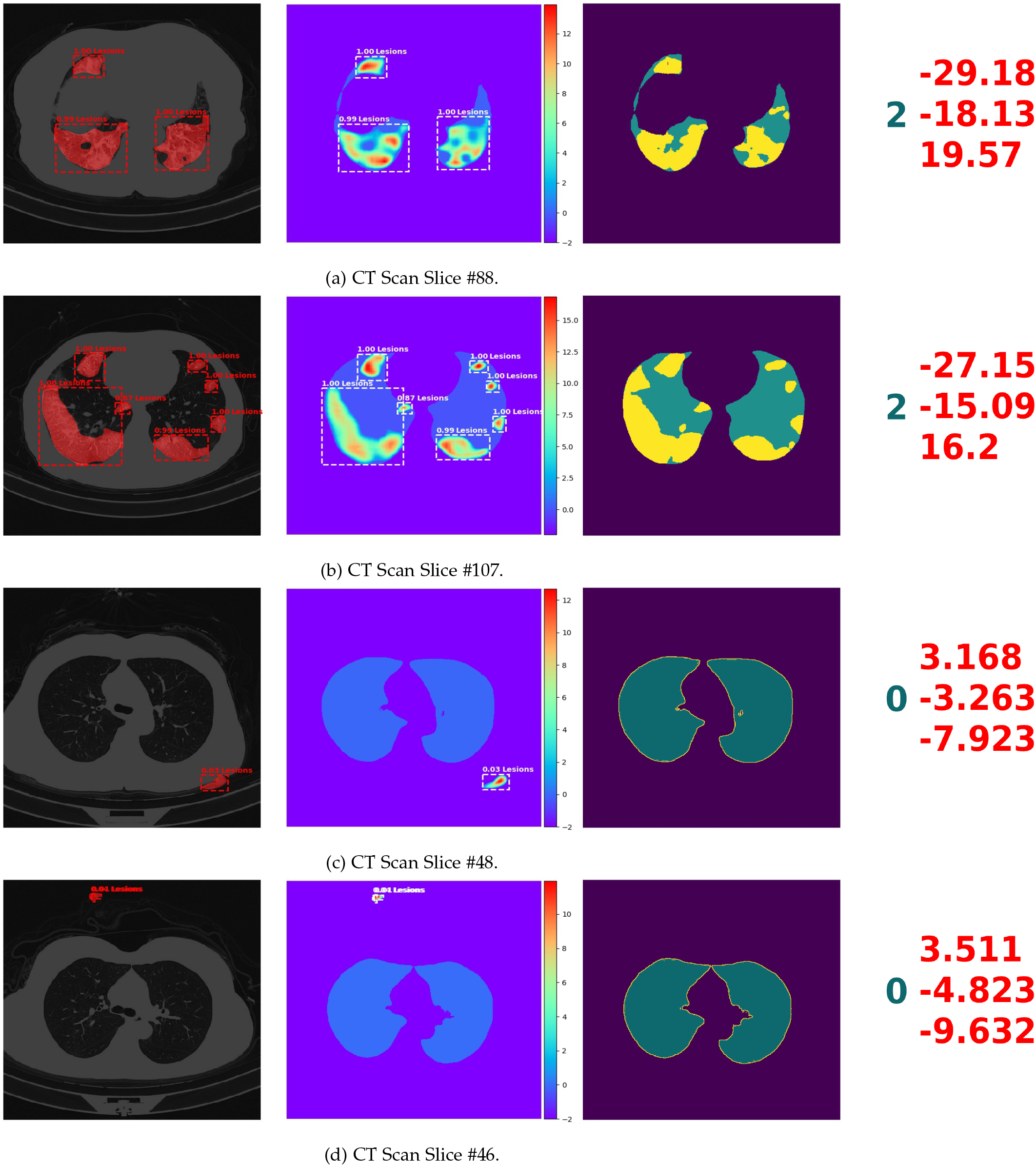
Segmentation results of ResNet34_*T*1_ model across a number of CT scan slices (different levels of the lungs). Images in each Figure 2a-2d pertain to the same scan slice. Figures 2a and 2b are COVID-19 positive, Figures 2c and 2d are Control/Negative. Column 1: Input images superimposed with the final mask prediction, bounding box, class and confidence scores for each instance. Column 2: Regional (mask) score maps. Outputs from each RoI are *independent of each other*, meaning that they were obtained from different RoIs independently and combined in the same score map. To avoid the image clutter, only the highest-ranking predictions are displayed. Column 3: Ground truth lesion and lungs masks. Column 4: true labels in dark green (0: Control, 2: COVID-19) and class scores predicted by COVID-CT-Mask-Net in red. Best viewed in color.

### 3.2 Distribution of observations in the RoI output batch

The analysis of the mask score maps in column 2, Figure 2 illustrates the effectiveness of the RoI batch to feature vector method, which is the main idea behind the transformation of Mask R-CNN into the classification model. Both the location (bounding box coordinates) and the importance (confidence score) of the areas critical to the COVID-19 diagnosis are output by RoI and accepted by the classification module **S** *in the decreasing order/rank of their importance/confidence scores*. Since the RoI batch size is fixed regardless of the RoIs’ confidence scores, **S** can learn this ranking, and, eventually, associate a number of RoIs located in the critical areas (see [ZZX^+^20, ZYW^+^20] for the analysis of COVID-19 vs Common Pneumonia chest CT scans) with the particular image class.

To demonstrate this, we plot the histograms of the confidence scores and the scatterplots of the confidence scores vs RoI area (bounding box size) in three difference CT scan slices, one for each class in Figure 3. Top 16 regions (columns 1-2) in Figure 3a are dominated by several mid-size (≈1000 pixels) high-scoring (≥ 0.95) critical areas, and the full batch (128 regions) in Columns 3-4 follows what seems to be a Exponential distribution. Therefore, despite the fact that the majority of regions have a very low score (regardless of the size), there is a sufficient number of high-scoring regions in the batch for the model to learn the true class. Common Pneumonia distribution is presented in Figure 3b: there’s a small number of mid to large (2000-4000 pixels) low to mid scoring regions with the scores between 0.1 and 0.3, but the majority of RoIs have a score close to 0. The distribution of Control(Negative), Figure 3b is also distinct: the highest-scoring box (0.001) is very large (≈ 8000 pixels), and the rest of the batch have scores practically indistinguishable from 0 regardless of the size.

**Figure 3:**
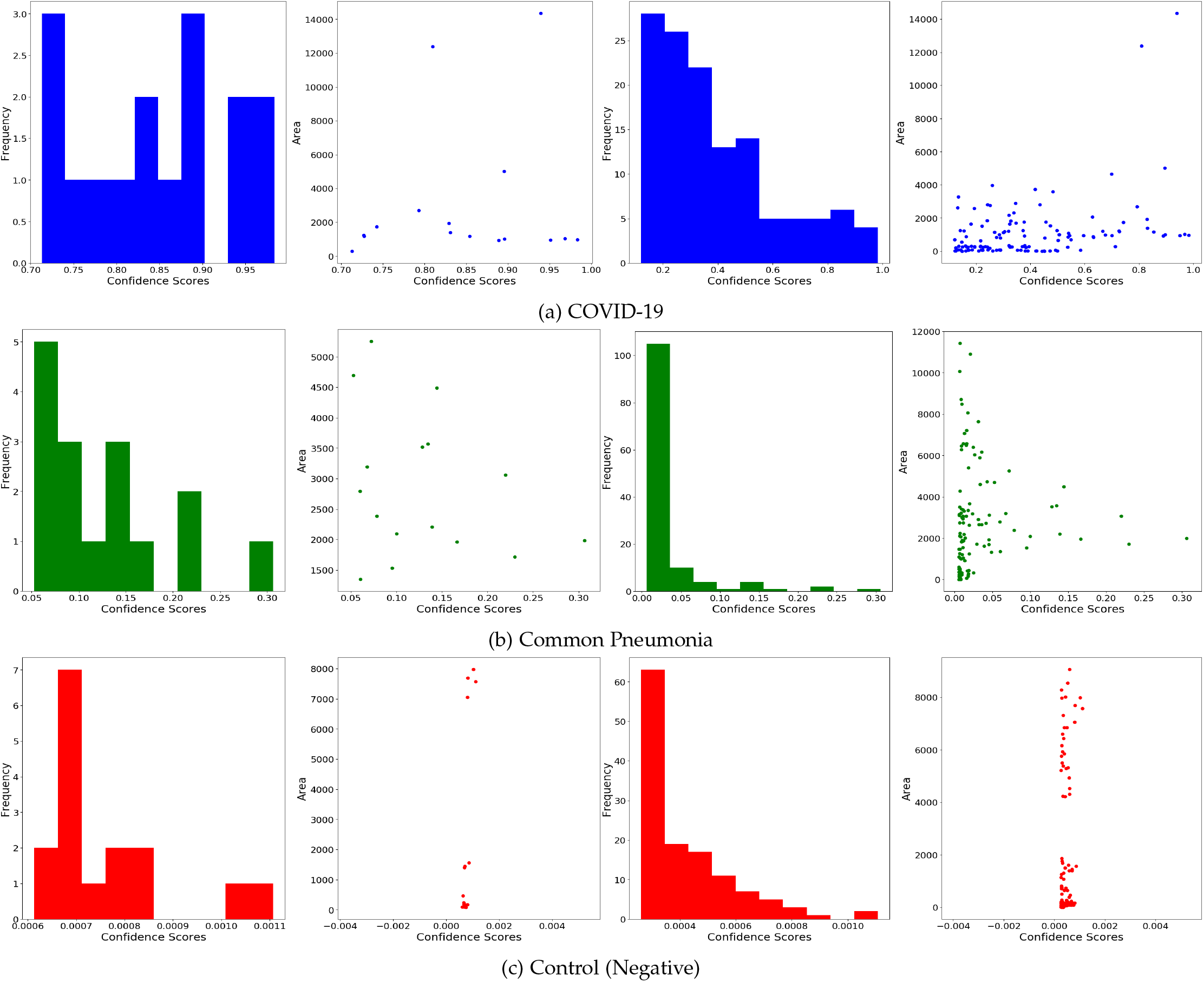
Distribution of the confidence score and scatterplot of the area vs the confidence score in a CT slice with COVID-19 (Figure 3a), Common pneumonia (Figure 3b) and Control (Figure 3c). Columns 1 and 2: top 16 predictions in each image, Columns 3 and 4: all 128 predictions in each image. Best viewed in color.

For a rigorous validation of the method, we also analyze the statistical distribution of the RoIs in the test sample (*n* = 1425) extracted from the test split, in which all 3 classes are equally represented (475 slices per class). We consider 3 RoI score_*θ*_:0.2, 0.5, 0.9. Each image is fed through COVID-CT-Mask-Net, and at the batch construction stage we extract the number of RoIs with confidence score exceeding these thresholds. Boxplots in Figure 4 and Table 4 present the mean and 95% confidence intervals (CI) for each class/threshold and Table 5 presents the results of one-sided Kolmogorov-Smirnov test comparing each pair of distribution at significance level *α* = 1%. This tests the hypothesis if the first distribution is less than the second and rejects it for p-values less than *α*. The only result that is not statistically significant is CP vs Normal, which explains a larger number of confusions and requires further investigation.

**Table 2:** Average precision of segmentation models and training time (in minutes). Best lightweight results in bold.

| Model | AP@0.5IoU | AP@0.75IoU | AP@[0.5 : 0.95] | Training |
| --- | --- | --- | --- | --- |
| ResNet18+FPN | 0.3699 | 0.2898 | 0.3137 | 128.75 |
| ResNet18<sub>T1</sub>+FPN | 0.4995 | 0.3778 | 0.3932 | 108.8 |
| ResNet18<sub>T2</sub>+FPN | 0.4993 | 0.3852 | 0.3759 | 126.9 |
| ResNet34+FPN | 0.5357 | 0.3333 | 0.3465 | 119.23 |
| ResNet34<sub>T1</sub>+FPN | **0.5988** | **0.4506** | **0.4476** | 134.01 |
| ResNet34<sub>T2</sub>+FPN | 0.4491 | 0.3118 | 0.3404 | 129.2 |
| ResNet50+FPN[TS20b](merged masks) | 0.6192 | 0.4522 | 0.4468 | 137.38 |
| ResNet50+FPN[TS20b](separate masks) | 0.5020 | 0.4198 | 0.3871 | 145.54 |

**Table 3:** Class sensitivity and overall accuracy results on COVIDx-CT test data (21192 images) and the training time (in minutes). Best lightweight results in bold.

| Model | COVID-19 | Pneumonia | Normal | Overall | Training |
| --- | --- | --- | --- | --- | --- |
| ResNet18+FPN | 87.51% | 77.31% | 74.57% | 78.18% | 151.2 |
| ResNet18<sub>T1</sub>+FPN | 91.35% | 91.63% | **96.98%** | **93.95%** | 106.6 |
| ResNet18<sub>T2</sub>+FPN | 84.05% | 85.81% | 93.01% | 88.66% | 113.66 |
| ResNet34+FPN | 86.98% | **94.27%** | 71.12% | 82.45% | 163.81 |
| ResNet34<sub>T1</sub>+FPN | **91.76%** | 91.70% | 94.36% | 92.89% | 144.73 |
| ResNet34<sub>T2</sub>+FPN | 89.25% | 93.32% | 92.11% | 91.99% | 112.6 |
| COVID-CT-Mask-Net<br>[TS20b](merged masks) | 92.68% | 96.69% | 97.74% | 96.63% | 430 |
| COVID-CT-Mask-Net<br>[TS20b](separate masks) | 93.88% | 95.06% | 96.91% | 95.64% | 460 |

**Table 4:**
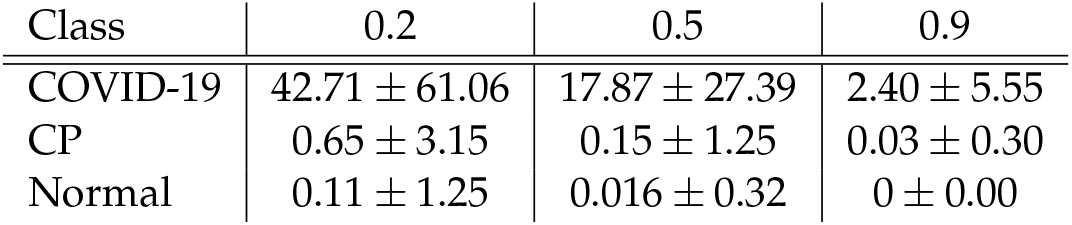
Mean number of RoIs exceeding the RoI score_*θ*_ for each class + 0.95% CI

**Table 5:** One-sided Kolmogorov-Smirnov Test Results for the scores exceeding the preset RoI score_*θ*_. Score/p-value. The result not statistically significant at *α* = 1% in bold.

| Class | 0.2 | 0.5 | 0.9 |
| --- | --- | --- | --- |
| COVID-19/CP | 0.96/1e-10 | 0.92/1e-10 | 0.63/1e-10 |
| COVID-19/Normal | 0.98/1e-10 | 0.94/1e-10 | N/A |
| CP/Normal | 0.18/2e-08 | **0.067/0.091** | N/A |

**Figure 4:**
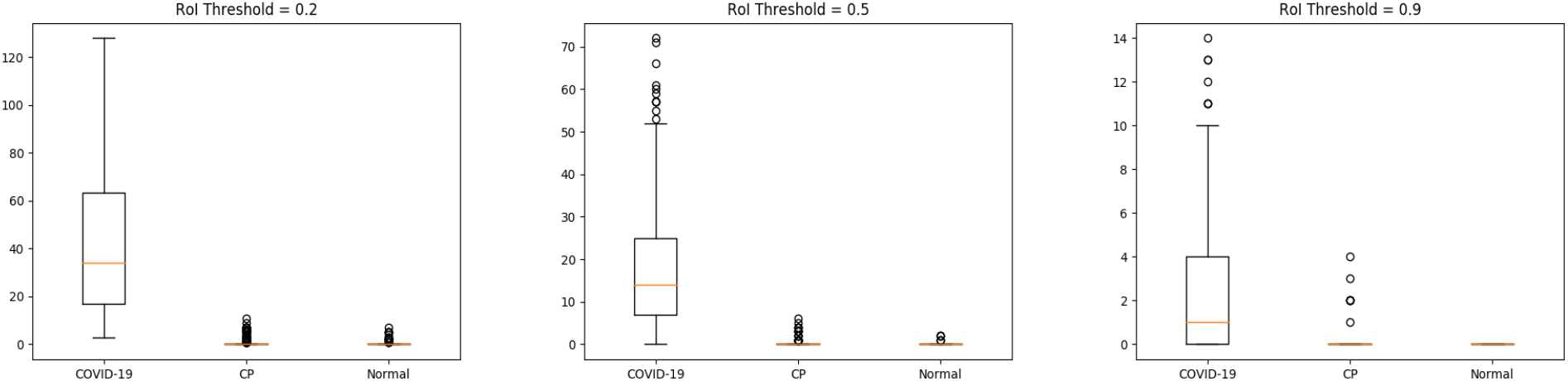
Boxplots of the number of RoIs for each class with scores*>*RoI score_*θ*_. Sample size:1425.

The value of this result is that, contrary to [LFBL20, ZYW^+^20], who showed that the differences in many COVID-19 and CP correlates are not statistically significant, the differences in the ranks of RoIs are mostly statistically significant across all 3 classes. Results in Figures 3 and 4 and Tables 4 and 5 were obtained with ResNet34_*T*1_+FPN.

## 4 Conclusions

We presented several variants of lightweight segmentation and classification models based on Mask R-CNN with ResNet18+FPN and ResNet34+FPN backbone networks. With as few as 11.74M total and 600K trainable parameters, COVID-CT-Mask-Net classification model with ResNet34_*T*1_+FPN backbone achieves a 91.76% COVID sensitivity and 92.89% overall accuracy across three classes (COVID-19, Common Pneumonia, Control). The model with ResNet18_*T*1_+FPN backbone with 6.35M parameters achieves the COVID-19 sensitivity of 91.35% and overall accuracy of 93.95%. The smallest model with ResNet18_*T*2_+FPN backbone with just 4.25M parameters achieves a 84.05% COVID-19 sensitivity and 88.66% overall accuracy. We also presented an in-depth analysis of the mask score maps across all three image classes and the distribution of the features of the predicted critical areas (confidence score, size). We demonstrated the ability of Mask R-CNN to explicitly detect and segment areas critical for the accurate prediction of COVID-19 and other classes.

## Data Availability

All the data is open-source and referenced accordingly in the paper.

https://github.com/AlexTS1980/COVID-CT-Mask-Net

